# Development and Validation of Machine Learning Models for Predicting Initiation of Emergency Dialysis in Advanced Chronic Kidney Disease

**DOI:** 10.64898/2026.06.21.26356128

**Authors:** Keita Hirano, Tomohisa Seki, Akihiro Watanabe, Kazumi Kubota, Yoshimasa Kawazoe

## Abstract

**Background:** Initiation of emergency dialysis, often requiring temporary catheter owing to unprepared definitive vascular access, is associated with infectious and vascular complications and suggests advanced chronic kidney disease (CKD) care gaps. Previous studies focused on kidney failure or dialysis timing. This study aimed to predict initiation of emergency dialysis using machine learning and baseline data.

**Methods:** This retrospective cohort study used the Japan Medical Data Center claims data (2014–2022). Adults with an estimated glomerular filtration rate (eGFR) <15 mL/min/1.73 m2 were included. The primary outcome was initiation of emergency dialysis (temporary catheter code without evidence of previous access preparation). Participants were randomly divided into derivation (80%) and validation (20%) cohorts. Logistic regression, support vector machine, XGBoost, LightGBM, and random forest models were evaluated using internal cross-validation, post-hoc calibration of the selected model, and bootstrap confidence intervals.

**Results:** The cohort included 3,062 individuals (derivation; n=2,449, validation; n=613). Emergency dialysis was initiated in 237 participants (7.7%); 185 (7.6%) and 52 (8.5%) in the derivation and validation cohorts, respectively. Validation area under the receiver operating characteristic curve ranged from 0.781–0.799, with the highest value observed for random forest (0.799, 95% confidence interval; 0.740–0.850). Risk stratification showed clear event enrichment in higher predicted risk categories. SHAP analyses identified hemoglobin, proteinuria, baseline eGFR, diabetes history, and diuretic use as key predictors. Decision curve analysis showed greater net benefit than eGFR alone at lower threshold probabilities.

**Conclusions:** Baseline machine learning models showed moderate discrimination for initiation of emergency dialysis and identified clinically plausible predictors. These findings support potential use for risk stratification, although external validation and evaluation within pre-specified care pathways are needed before implementation.

## Introduction

Chronic kidney disease (CKD) is common and progressive, often leading to dialysis.^1,2^ Dialysis initiation is either planned, in which vascular access is prepared in advance, or emergency, in which dialysis is initiated with temporary catheter insertion because access is not yet available. Emergency initiation is associated with higher risks of infection and vascular complications, early death, and rehospitalization.^3^ It imposes substantial burdens on quality of life and healthcare costs, whereas planned initiation with a functioning arteriovenous fistula and adequate predialysis nephrology care is associated with better outcomes.^4–10^ Previous studies have emphasized that unplanned initiation is clinically heterogeneous and may reflect broader gaps in CKD care pathways.^11–15^

Existing research has mainly focused on kidney failure or the timing of dialysis initiation. Representative kidney failure prediction equations rely on a small set of routinely available variables and have been evaluated across multiple cohorts and countries.^16–19^ Other models have incorporated longitudinal information, such as eGFR trajectories, or have targeted renal replacement therapy within medium-term horizons to support referral and preparation timing.^20–22^ However, these frameworks treat kidney failure or renal replacement therapy as the outcome and do not explicitly distinguish planned from emergency initiation. Consequently, even a well-performing kidney failure model may have limited value if the clinical question is whether a patient may start dialysis under suboptimal circumstances, in which potentially preventable steps, such as timely access creation, can still be conducted.

Recently, machine learning methods have been applied to predict dialysis initiation or kidney failure within fixed time windows, often with high discrimination.^23–30^ Although unplanned dialysis is discussed, operational definitions vary across settings, and the models are often difficult to translate into practice without clear linkage to modifiable clinical actions. Recent research has begun to address this gap by developing machine learning models intended to prevent unplanned dialysis,^31^ but the target outcome in many studies remains near-term kidney failure. An updated systematic review summarized the risk factors for unplanned initiation, including older age, diabetes, poor nutritional status, and delayed nephrology referral. However, these factors have rarely been integrated into a dedicated prediction model focused specifically on emergency initiation as the primary outcome.^32^

Accordingly, this study aimed to develop and validate machine learning models for initiation of emergency dialysis using routinely available baseline health checkups and claims data. Emergency initiation was defined as dialysis that started with temporary catheter insertion without previous access preparation. By focusing directly on this clinically consequential transition, we sought to evaluate whether high-risk patients could be identified sufficiently early to inform intensified follow-up and access planning.

## Methods

### Study design and data sources

This retrospective cohort study used data from the Japan Medical Data Center (JMDC) insurer-based claims database between January 2014 and December 2022. The JMDC database integrates subscriber registries from employment-based health insurance societies with claims generated across medical institutions and results from annual health checkups. Because claims are centrally managed under Japan’s universal coverage system, the database allows longitudinal tracking of diagnoses and treatments across healthcare facilities unless an individual changes or withdraws from the insurer.

We constructed a cohort of adults with advanced CKD by linking health checkup data with claims-based records, including procedures, prescriptions, diagnoses, and medical materials. Baseline was defined as the first health check-up record at which the estimated glomerular filtration rate (eGFR) was <15 mL/min/1.73 m2, calculated using the Japanese coefficient-modified serum creatinine equation. Participants with previous dialysis, follow-up periods <180 days, or serum creatinine values considered clinically implausible and likely to reflect data entry or unit-recording errors were excluded. After applying these criteria, the final analytical cohort comprised 3,062 individuals (Figure 1). Follow-up began on the baseline date and continued until the earliest initiation of emergency dialysis, death, insurance disenrollment, or end of data availability.

**Figure 1.**
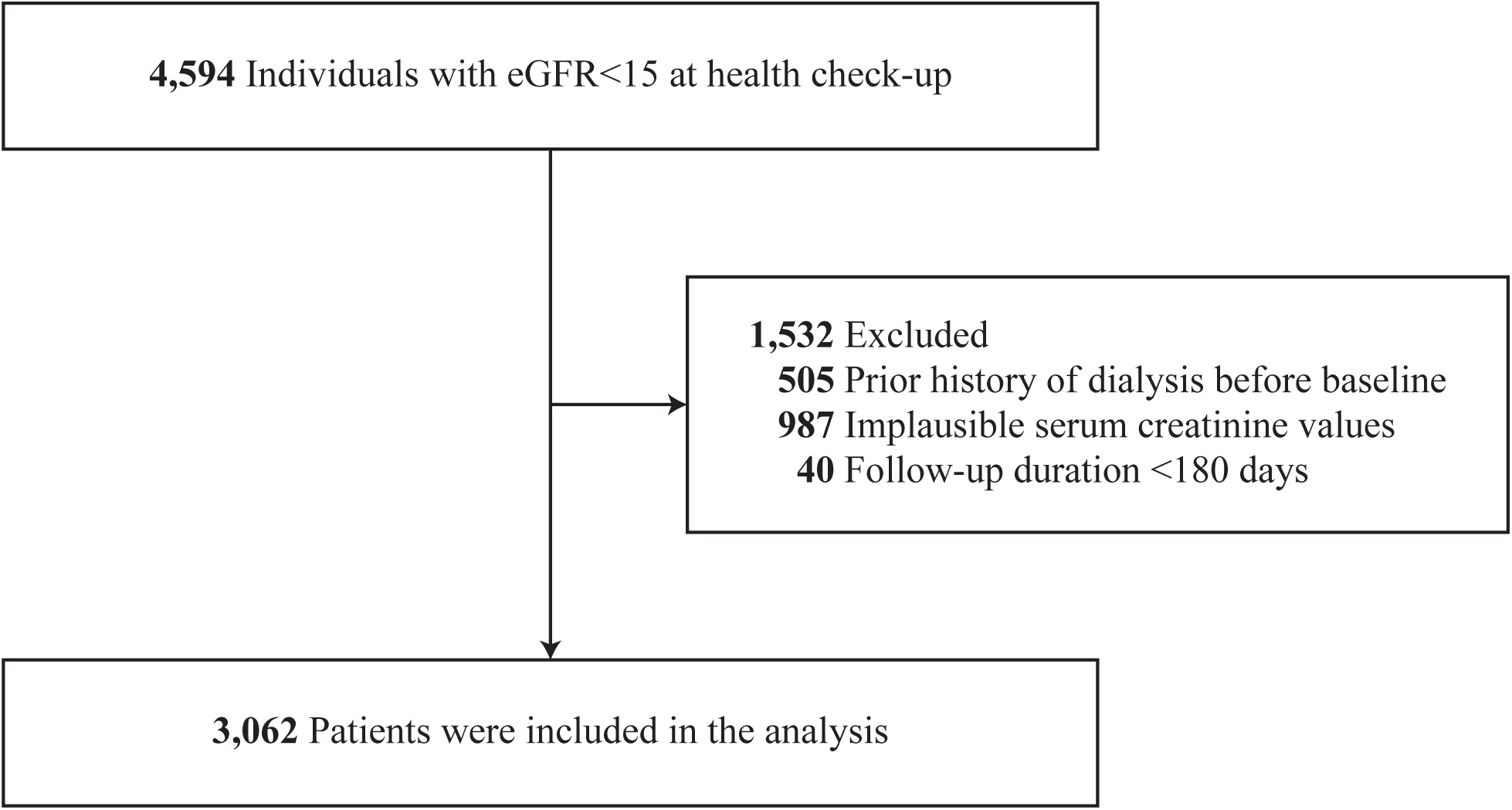
Study Flow Diagram Flow diagram of cohort assembly from the source population to the final analytic cohort, including the exclusion criteria. Abbreviations: eGFR, estimated glomerular filtration rate.

**Figure 2.**
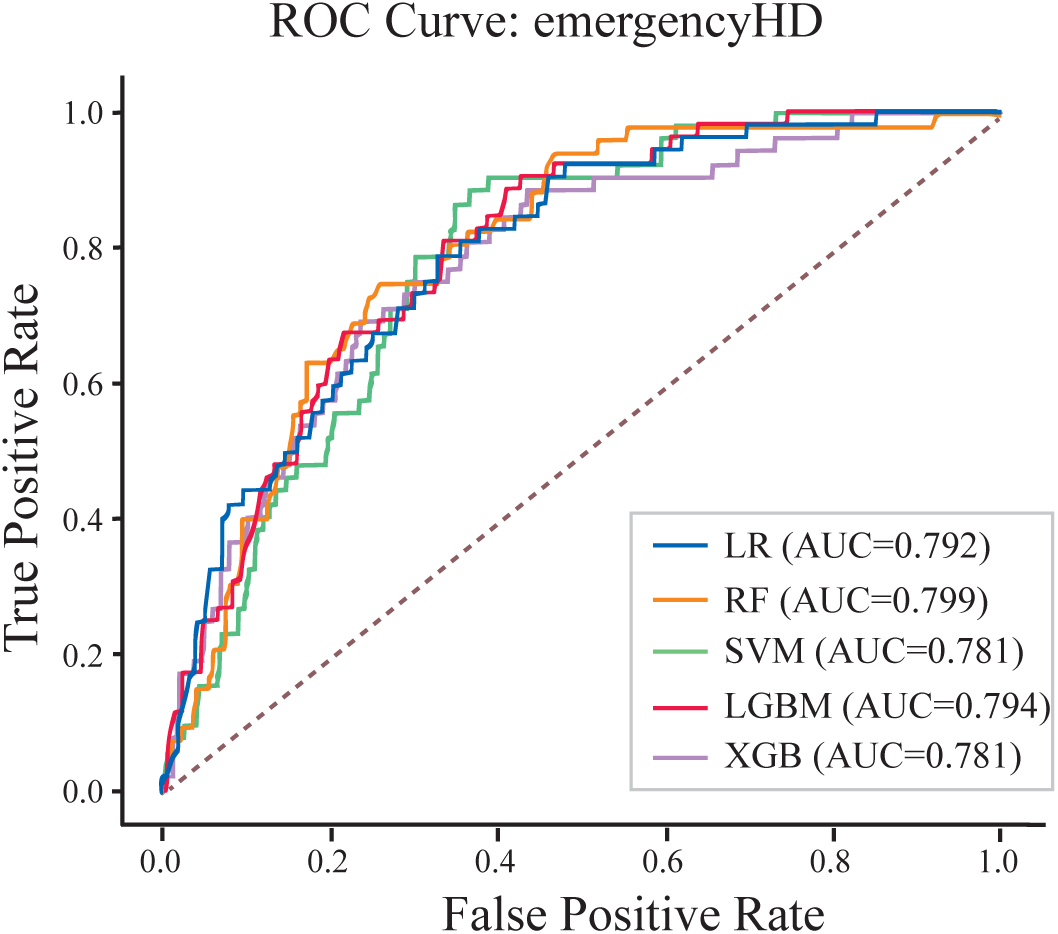
Receiver Operating Characteristic Curves for Predicting Initiation of Emergency Dialysis in the Validation Cohort Receiver operating characteristic curves for each model in the validation cohort. C-statistics are shown for each model. Abbreviations: LR, logistic regression; RF, random forest; SVM, support vector machine; LGBM, Light Gradient Boosting Machine; XGB, extreme gradient boosting; ROC, receiver operating characteristic.

**Figure 3.**
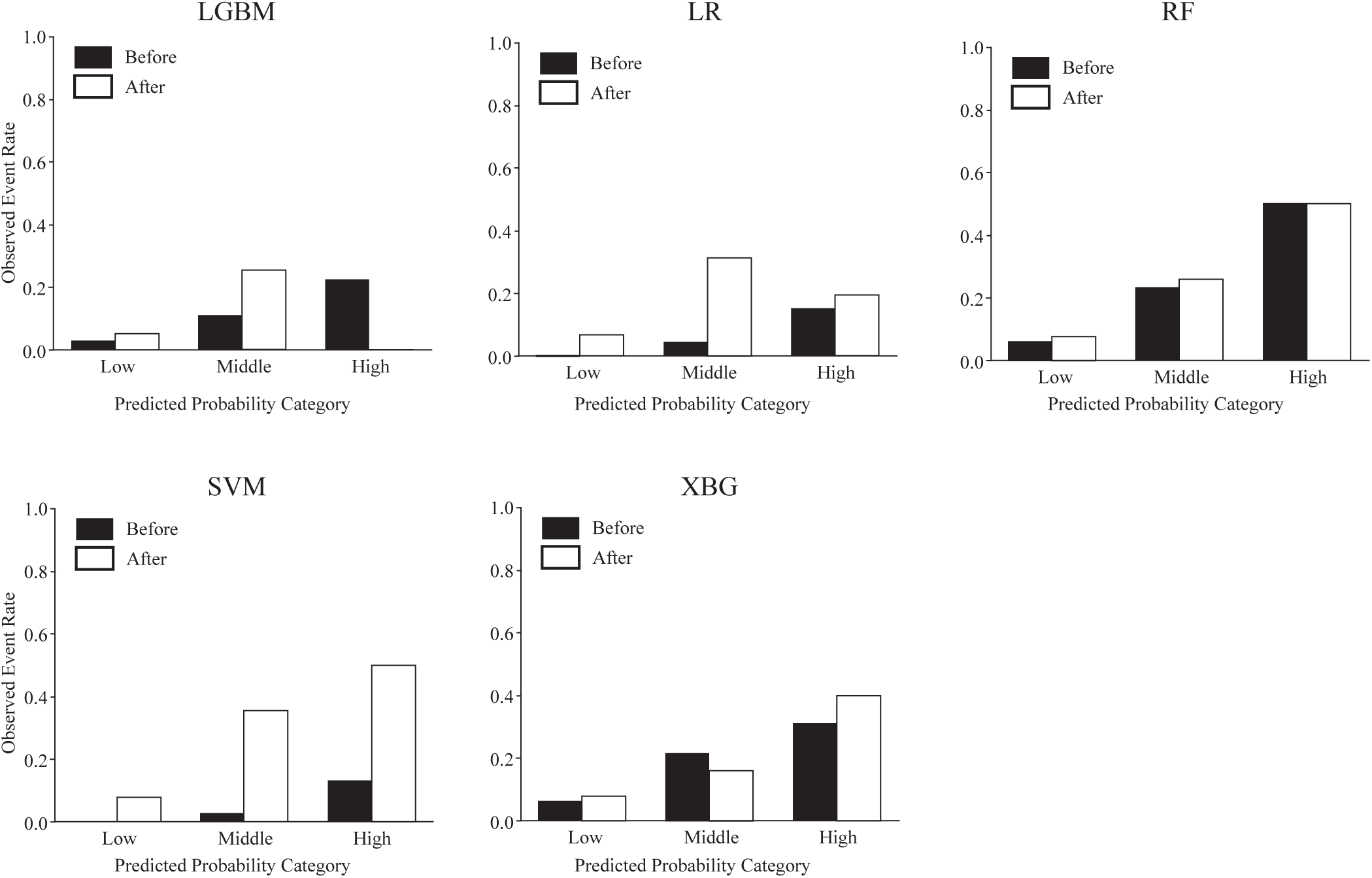
Risk-Stratified Calibration Before and After Recalibration in the Validation Cohort Observed event rates across risk categories before and after recalibration. Abbreviations: LR, logistic regression; RF, random forest; SVM, support vector machine; LGBM, Light Gradient Boosting Machine; XGB, extreme gradient boosting.

### Study population

We included adult subscribers who underwent health check-ups during the study period and had advanced CKD. The baseline date was the date of the first health checkup to meet the eGFR threshold. The exclusion criteria were dialysis procedure codes before baseline, follow-up <180 days, and serum creatinine values considered clinically implausible and likely to reflect data entry or unit-recording errors. Participants were followed from baseline until the earliest initiation of emergency dialysis, death, loss of insurance eligibility, or the end of the observation window. The study size was determined by the number of eligible individuals in the JMDC database who met the inclusion and exclusion criteria during the study period.

### Predictors

Candidate predictors were defined based on information from health check-up or claim-derived records available before baseline. Demographic variables included age and sex. Anthropometric and vital sign variables included body mass index (BMI), systolic BP, and diastolic BP. Laboratory variables included serum creatinine (Cr), estimated glomerular filtration rate (eGFR), hemoglobin (Hgb), aspartate aminotransferase (AST), alanine aminotransferase (ALT), gamma-glutamyl transpeptidase (GGT), LDL cholesterol, HDL cholesterol, triglycerides (TG), fasting blood sugar (FBS), and hemoglobin A1c (HbA1c). Urinalysis variables included hematuria and proteinuria categories. Lifestyle-related factors included current smoking, alcohol consumption, poor sleep, subjective symptoms, and objective symptoms.

Comorbidities identified prior to baseline included hypertension, diabetes, dyslipidemia, cardiovascular disease, cerebrovascular disease, acute kidney injury, and anemia. Medication use was defined based on prescription records prior to the baseline assessment and included antihypertensive drugs, antidiabetic drugs, lipid-lowering drugs, potassium binders, and diuretics. Missing values were imputed within leak-free preprocessing pipelines to avoid information leakage from the validation cohort.

### Outcomes

The primary outcome was initiation of emergency dialysis, defined as the presence of an emergency blood access catheter material code without any record of shunt creation or permanent vascular access within 30 days of the first dialysis. The modeling target was the initiation of emergency dialysis at any time during the available follow-up after cohort entry. Participants without initiation of emergency dialysis were classified as non-events; this group included individuals who later initiated dialysis in a planned fashion and those who did not initiate dialysis during the observed follow-up. This framework was selected because the intended use of the model was baseline risk stratification among all patients with advanced CKD at the time they were first identified. Secondary outcomes included dialysis initiation within 90 days, one year, and three years from the baseline, each defined as a binary outcome.

### Statistical analysis

The baseline characteristics were summarized descriptively. Participants were randomly assigned to the derivation (80%) and validation (20%) cohorts using a fixed random seed. Candidate algorithms included logistic regression, random forest, support vector machine (SVM), extreme gradient boosting (XGBoost), and Light Gradient Boosting Machine (LightGBM). All preprocessing steps, including imputation, encoding, standardization (as required), and class-imbalance handling, were implemented within leak-free pipelines. The synthetic minority oversampling technique was applied only within the derivation data as appropriate; if event counts were too sparse, class weighting was used. For the random forest model, the hyperparameters were optimized within the derivation cohort using Optuna with a Tree-structured Parzen Estimator sampler. The search space included the number of trees (300–1500), maximum depth (3–30), minimum number of samples per leaf (1–10), and maximum feature parameters (sqrt, log2, or none), with the average precision as the optimization objective in internal cross-validation. The model performance was evaluated using the area under the receiver operating characteristic (ROC) curve (AUC), area under the precision-recall curve, accuracy, precision, recall, F1 score, and Matthews correlation coefficient. For the selected model, post-hoc probability calibration was assessed within the derivation cohort using isotonic regression if class counts were permitted, and sigmoid calibration otherwise; calibrated probabilities were evaluated in the validation cohort. Confidence intervals (CIs) were estimated by bootstrap resampling. Calibration, confusion matrices, SHAP-based feature-contribution analyses, and decision curve analyses were performed as supportive evaluations. For decision curve analysis, net benefit was calculated in the validation cohort across prespecified threshold probabilities for the selected model, treat-all strategy, treat-none strategy, and an eGFR-alone logistic regression model. All analyses were performed using the Python version 3.8.8 and R version 4.0.2 (R Foundation for Statistical Computing, Vienna, Austria).

### Ethics Approval

This study was approved by the Ethics Committee of the University of Tokyo Hospital (Approval No. 2024103NIe) and was conducted in accordance with the Declaration of Helsinki. The requirement for informed consent was waived because this study used anonymized claims and health check-up data. Reporting followed the STROBE guidelines for observational studies.

## Results

### Study enrollment and cohort stratification

The analysis involved 3,062 individuals, randomly divided into derivation (n = 2,449) and validation (n = 613) cohorts. Emergency dialysis was initiated in 237 patients (7.7%);185 (7.6%) and 52 (8.5%) in the derivation and validation cohorts, respectively (Table 1). The patients’ median age was 53.0 years (IQR; 47.0–59.0), and 81.7% were men. Median baseline eGFR was 7.38 mL/min/1.73 m2 (IQR; 4.92–12.08). Although the split was random, several baseline characteristics differed between cohorts, including diabetes prevalence, diuretic use, and eGFR, reflecting sampling variability in a relatively small, high-risk cohort.

**Table 1.**
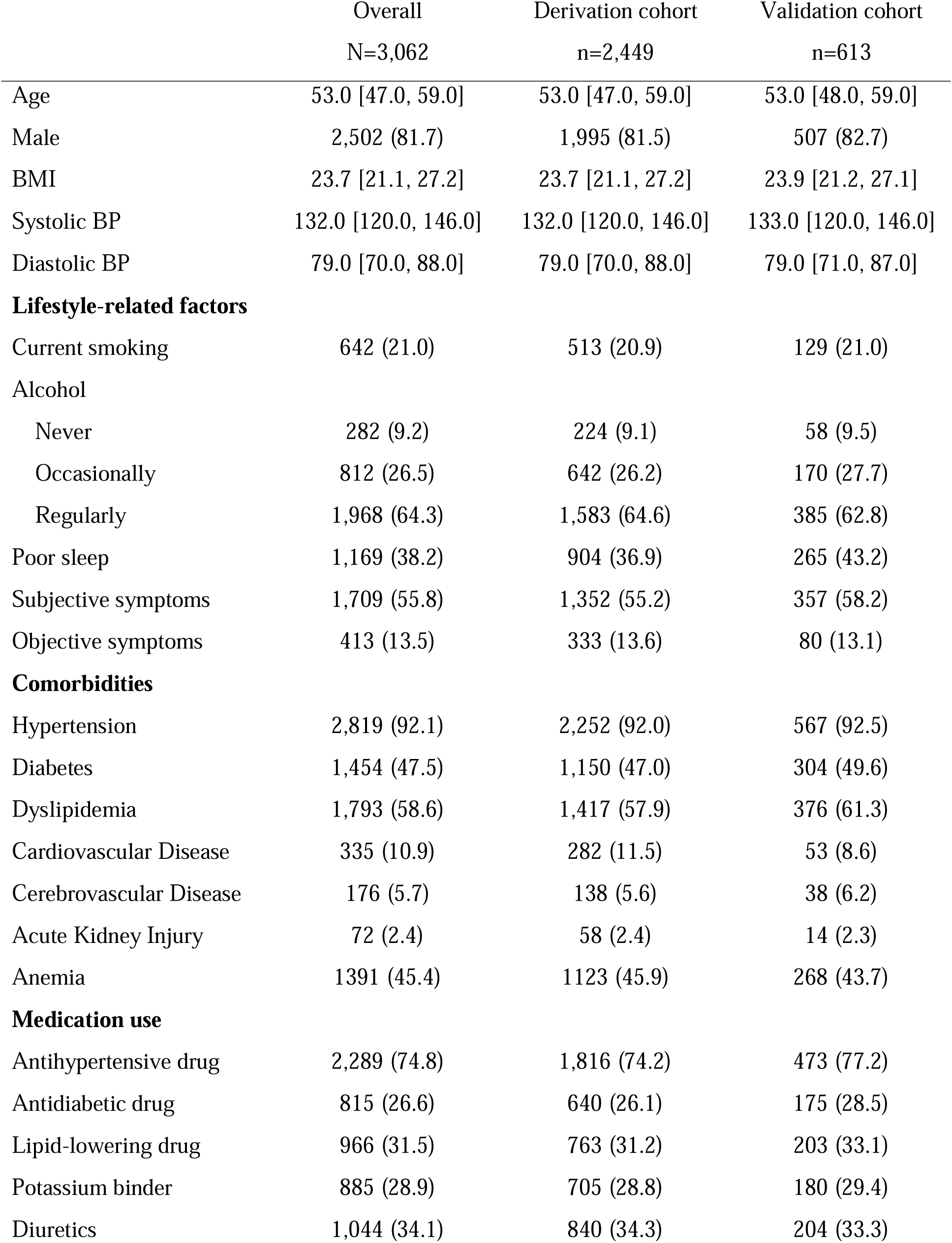
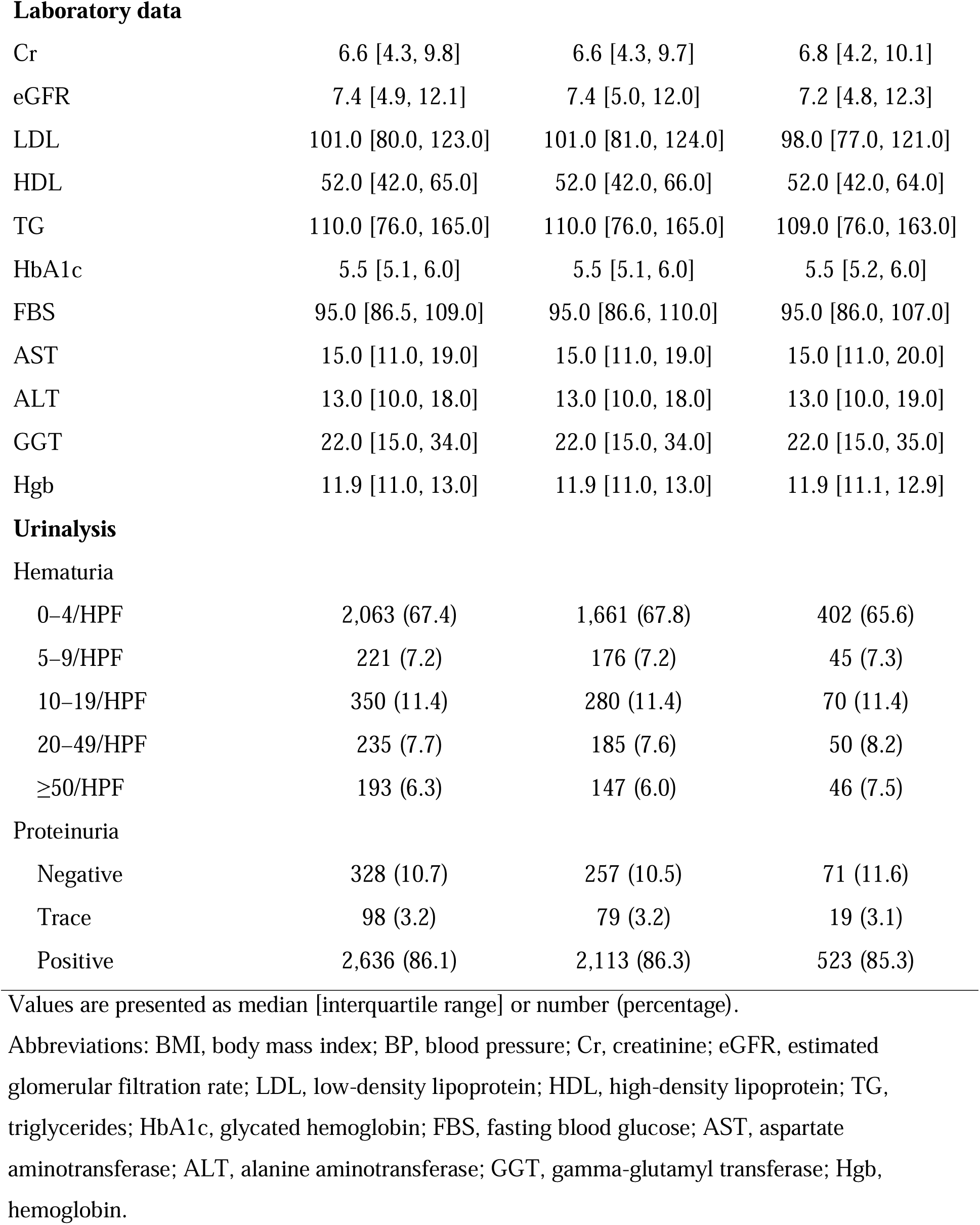
Baseline characteristics of the derivation and validation cohorts.

|  | Overall<br>N=3,062 | Derivation cohort<br>n=2,449 | Validation cohort<br>n=613 |
| --- | --- | --- | --- |
| Age | 53.0 [47.0, 59.0] | 53.0 [47.0, 59.0] | 53.0 [48.0, 59.0] |
| Male | 2,502 (81.7) | 1,995 (81.5) | 507 (82.7) |
| BMI | 23.7 [21.1, 27.2] | 23.7 [21.1, 27.2] | 23.9 [21.2, 27.1] |
| Systolic BP | 132.0 [120.0, 146.0] | 132.0 [120.0, 146.0] | 133.0 [120.0, 146.0] |
| Diastolic BP | 79.0 [70.0, 88.0] | 79.0 [70.0, 88.0] | 79.0 [71.0, 87.0] |
| **Lifestyle-related factors** |  |  |  |
| Current smoking | 642 (21.0) | 513 (20.9) | 129 (21.0) |
| Alcohol |  |  |  |
| Never | 282 (9.2) | 224 (9.1) | 58 (9.5) |
| Occasionally | 812 (26.5) | 642 (26.2) | 170 (27.7) |
| Regularly | 1,968 (64.3) | 1,583 (64.6) | 385 (62.8) |
| Poor sleep | 1,169 (38.2) | 904 (36.9) | 265 (43.2) |
| Subjective symptoms | 1,709 (55.8) | 1,352 (55.2) | 357 (58.2) |
| Objective symptoms | 413 (13.5) | 333 (13.6) | 80 (13.1) |
| **Comorbidities** |  |  |  |
| Hypertension | 2,819 (92.1) | 2,252 (92.0) | 567 (92.5) |
| Diabetes | 1,454 (47.5) | 1,150 (47.0) | 304 (49.6) |
| Dyslipidemia | 1,793 (58.6) | 1,417 (57.9) | 376 (61.3) |
| Cardiovascular Disease | 335 (10.9) | 282 (11.5) | 53 (8.6) |
| Cerebrovascular Disease | 176 (5.7) | 138 (5.6) | 38 (6.2) |
| Acute Kidney Injury | 72 (2.4) | 58 (2.4) | 14 (2.3) |
| Anemia | 1391 (45.4) | 1123 (45.9) | 268 (43.7) |
| **Medication use** |  |  |  |
| Antihypertensive drug | 2,289 (74.8) | 1,816 (74.2) | 473 (77.2) |
| Antidiabetic drug | 815 (26.6) | 640 (26.1) | 175 (28.5) |
| Lipid-lowering drug | 966 (31.5) | 763 (31.2) | 203 (33.1) |
| Potassium binder | 885 (28.9) | 705 (28.8) | 180 (29.4) |
| Diuretics | 1,044 (34.1) | 840 (34.3) | 204 (33.3) |
| **Laboratory data** |  |  |  |
| Cr | 6.6 [4.3, 9.8] | 6.6 [4.3, 9.7] | 6.8 [4.2, 10.1] |
| eGFR | 7.4 [4.9, 12.1] | 7.4 [5.0, 12.0] | 7.2 [4.8, 12.3] |
| LDL | 101.0 [80.0, 123.0] | 101.0 [81.0, 124.0] | 98.0 [77.0, 121.0] |
| HDL | 52.0 [42.0, 65.0] | 52.0 [42.0, 66.0] | 52.0 [42.0, 64.0] |
| TG | 110.0 [76.0, 165.0] | 110.0 [76.0, 165.0] | 109.0 [76.0, 163.0] |
| HbA1c | 5.5 [5.1, 6.0] | 5.5 [5.1, 6.0] | 5.5 [5.2, 6.0] |
| FBS | 95.0 [86.5, 109.0] | 95.0 [86.6, 110.0] | 95.0 [86.0, 107.0] |
| AST | 15.0 [11.0, 19.0] | 15.0 [11.0, 19.0] | 15.0 [11.0, 20.0] |
| ALT | 13.0 [10.0, 18.0] | 13.0 [10.0, 18.0] | 13.0 [10.0, 19.0] |
| GGT | 22.0 [15.0, 34.0] | 22.0 [15.0, 34.0] | 22.0 [15.0, 35.0] |
| Hgb | 11.9 [11.0, 13.0] | 11.9 [11.0, 13.0] | 11.9 [11.1, 12.9] |
| Hematuria |  |  |  |
| 0–4/HPF | 2,063 (67.4) | 1,661 (67.8) | 402 (65.6) |
| 5–9/HPF | 221 (7.2) | 176 (7.2) | 45 (7.3) |
| 10–19/HPF | 350 (11.4) | 280 (11.4) | 70 (11.4) |
| 20–49/HPF | 235 (7.7) | 185 (7.6) | 50 (8.2) |
| ≥50/HPF | 193 (6.3) | 147 (6.0) | 46 (7.5) |
| Proteinuria |  |  |  |
| Negative | 328 (10.7) | 257 (10.5) | 71 (11.6) |
| Trace | 98 (3.2) | 79 (3.2) | 19 (3.1) |
| Positive | 2,636 (86.1) | 2,113 (86.3) | 523 (85.3) |

Values are presented as median [interquartile range] or number (percentage).
Abbreviations: BMI, body mass index; BP, blood pressure; Cr, creatinine; eGFR, estimated glomerular filtration rate; LDL, low-density lipoprotein; HDL, high-density lipoprotein; TG, triglycerides; HbA1c, glycated hemoglobin; FBS, fasting blood glucose; AST, aspartate aminotransferase; ALT, alanine aminotransferase; GGT, gamma-glutamyl transferase; Hgb, hemoglobin.

### Model performance and calibration

The discrimination for initiation of emergency dialysis varied across algorithms. In the validation cohort, AUC values were 0.792, 0.799, 0.781, 0.794, and 0.781 for logistic regression, random forest, SVM, LightGBM, and XGBoost, respectively (Table 2). The random forest model showed the highest validation AUC and was examined in greater detail. The risk-stratified calibration of the random forest model showed increasing observed event rates across the low-, middle-, and high-risk groups. Before calibration, the observed event rates were 0.017, 0.052, and 0.427, respectively; after calibration and reclassification using calibrated probabilities, the corresponding values were 0.028, 0.063, and 0.667. Thus, although the overall discrimination was moderate, the model concentrated on events within the highest risk category. Additional performance metrics, including the accuracy, precision, recall, F1 score, and Matthews correlation coefficient, are presented in Supplementary Table 1.

**Table 2.** C-statistics of different models in the derivation and validation cohort.

|  | Derivation cohort | Validation cohort |
| --- | --- | --- |
| LR | 0.749 (0.719-0.780) | 0.792 (0.736-0.851) |
| RF | 0.752 (0.721-0.785) | **0.799 (0.740-0.850)** |
| SVM | 0.736 (0.704-0.768) | 0.781 (0.721-0.842) |
| LGBM | 0.774 (0.744-0.800) | 0.794 (0.742-0.844) |
| XGB | 0.745 (0.713-0.776) | 0.781 (0.727-0.830) |

Values are presented as c-statistics (95% confidence intervals). Abbreviations: LR, logistic regression; RF, random forest; SVM, support vector machine; LGBM, Light Gradient Boosting Machine; XGB, extreme gradient boosting.

### SHAP value

Feature importance assessed by SHAP values indicated that eGFR, hemoglobin, proteinuria, smoking, antihypertensive medication use, poor sleep, serum creatinine, anemia, history of cardiovascular disease, and diabetes were among the strongest contributors to model output (Figure 4). The relative ranking of these variables supports the model’s validity, although the SHAP values should be interpreted as indicators of predictive contribution.

**Figure 4.**
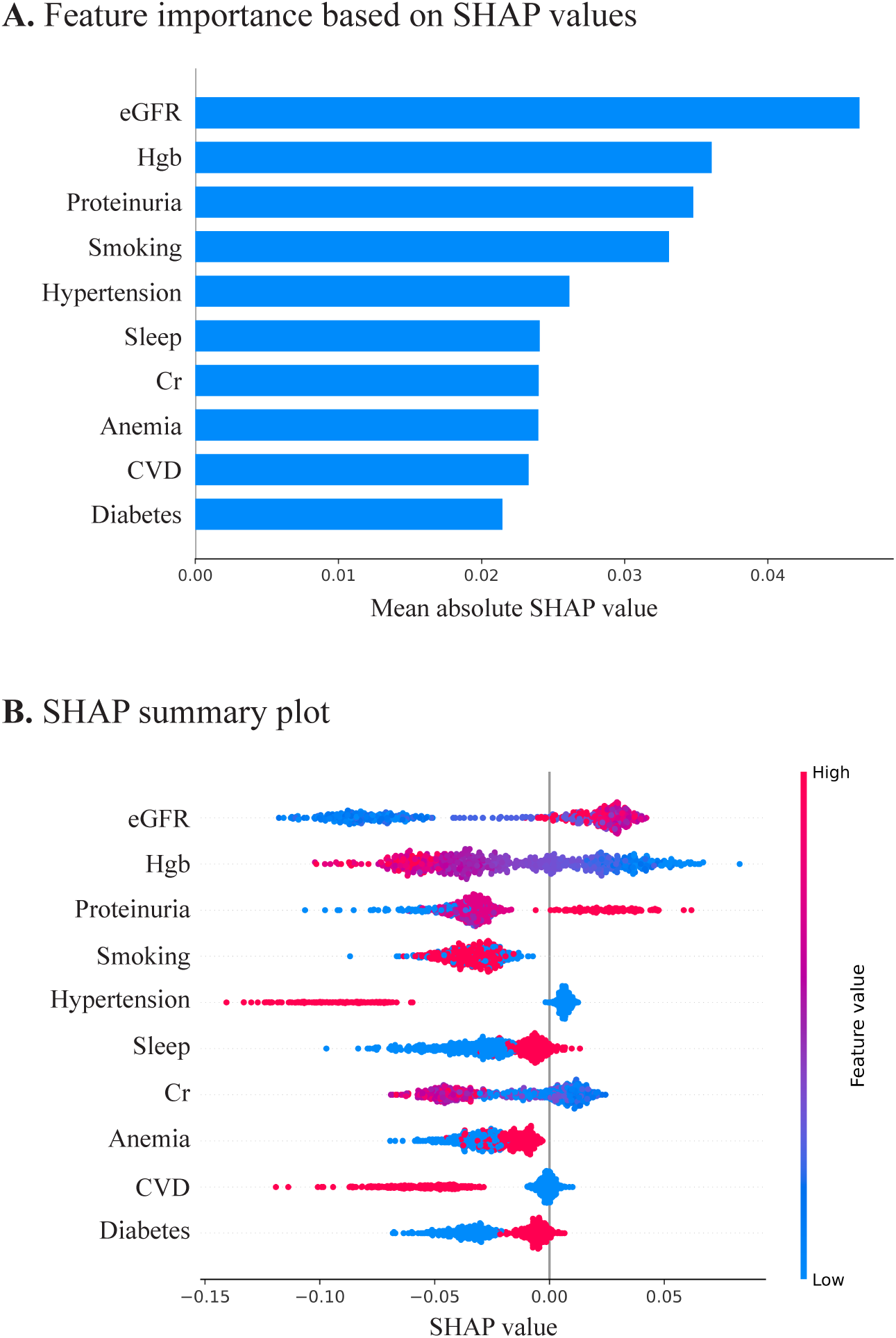
Contributions of Baseline Characteristics in the Prediction Model for the Initiation of Emergency Dialysis SHAP, Shapley additive explanations. Mean absolute SHAP values represent the overall contribution of each feature to model prediction. In the SHAP summary plot, positive SHAP values indicate higher predicted risk, whereas negative values indicate lower predicted risk. Colors indicate feature values, with red and blue representing higher and lower values, respectively.

**Figure 5.**
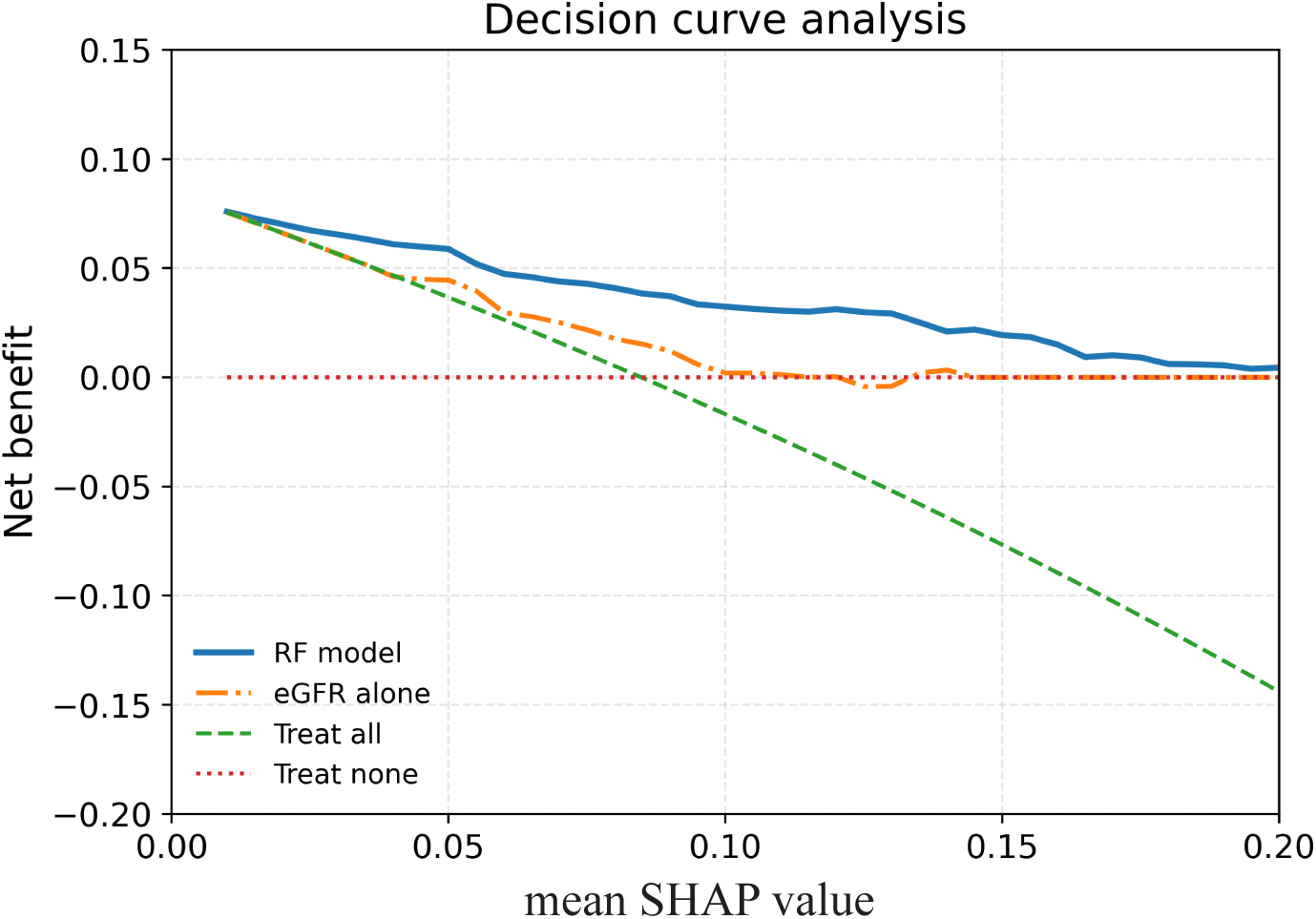
Clinical Utility of the Random Forest Model for Predicting the Initiation of Emergency Dialysis Net benefit across threshold probabilities compared with eGFR alone, treat-all, and treat-none strategies. The random forest model was selected as the best-performing model based on the C-statistics.

### Decision curve analysis

Decision curve analysis showed that the selected model provided greater net benefit than eGFR alone at lower threshold probabilities. The model also showed higher net benefit than the treat-all and treat-none strategies across most of this range.

## Discussion

Initiation of emergency dialysis remains a persistent and clinically important problem in the care of patients with advanced CKD. Patients who start dialysis urgently, typically with a temporary catheter and often during hospitalization, experience worse outcomes than those who transition in a planned manner. Previous studies have shown that unplanned initiation remains common despite improvements in pre-dialysis care.^32^ This study developed machine learning models to predict initiation of emergency dialysis and found that a random forest model achieved the best performance (AUC; 0.799). The model demonstrated reasonable calibration for risk grouping and clear enrichment of events in the higher-predicted-risk strata.

Existing prediction tools address different clinical questions. The Kidney Failure Risk Equation and related models estimate the risk of kidney failure over defined time horizons and have been widely validated across populations.^17^ Recent approaches have extended this paradigm by incorporating competing risks and estimating the timing of kidney failure and related outcomes.^20^ However, these models do not directly address how dialysis is initiated. Even recent short-term prediction models designed to reduce unplanned dialysis primarily target near-term kidney failure.^31^ Contrastingly, this study focuses on a distinct clinical endpoint; failure of a safe transition to dialysis. This distinction is critical because the clinical challenge in advanced CKD includes ensuring that transition occurs in a planned and controlled manner.

This difference explains that initiation of emergency dialysis is inherently more difficult to predict. Observational studies consistently show that urgent initiation arises from illness-, service-, and patient-related factors, including acute decompensation, delayed access planning, and limited engagement in pre-dialysis care.^3,15^ Clinical guidelines reinforce that dialysis initiation should be based on eGFR and a broader clinical assessment, including symptoms, metabolic complications, and overall condition.^1,2^ Collectively, these findings suggest that emergency initiation reflects disease trajectory and the success or failure of care pathways. In this setting, an AUC of 0.799 is clinically meaningful, as the model attempts to predict a complex care-transition event.

The pattern of the model predictors is consistent with this interpretation. eGFR was the strongest contributor, but was accompanied by hemoglobin, proteinuria, diabetes, cardiovascular disease, and diuretic use, suggesting that the model captured a broader phenotype of instability. This aligns with previous literature indicating that anemia, comorbidity burden, and markers of disease progression are associated with unplanned dialysis initiation.^32^ Although a lower eGFR may be associated with dialysis initiation, our results suggest that some patients with a relatively preserved eGFR still undergo emergency initiation. Clinically, patients with less severely reduced eGFR may not trigger timely access planning or preparation yet may deteriorate abruptly because of volume overload, infection, or cardiovascular events. Thus, the key message is that reliance on kidney function alone may underestimate transition risk.

The potential role of this model lies in supporting the earlier identification of high-risk patients rather than replacing clinical decision-making. Prediction models in nephrology are increasingly expected to inform care pathways.^23,26^ In this study, the decision curve analysis suggested that the model may have value beyond eGFR alone at lower threshold probabilities relevant to screening and early intervention. This supports the use of a triage or early warning tool to prompt earlier vascular access planning, repeated modality education, and closer follow-up, rather than as a stand-alone trigger for treatment decisions. This interpretation is consistent with current trends in clinical prediction modeling, in which modest improvements in early identification may translate into meaningful improvements in care processes.

The superior performance of random forest is consistent with previous evidence that tree-based methods perform well in structured, medium-sized clinical datasets, in which nonlinear relationships and heterogeneous features are common. The observed performance is methodologically plausible and does not necessarily imply that more complex models, such as deep learning, would provide additional benefits.

This study had several limitations. First, the outcome was modeled as a binary event over the available follow-up period rather than as a time-to-event outcome, and competing risks, such as death, were not explicitly modeled. Second, the non-event group included heterogeneous patients, including those who initiated dialysis in a planned manner and those who did not initiate dialysis, reflecting the intended baseline risk stratification, but limiting interpretation as a direct comparison between emergency and planned initiation. Third, the outcome definition relied on claims-based codes and may not fully capture access preparation, patient preference, or the acute clinical context. Fourth, the validation was internal, and external validation in independent cohorts was essential. Fifth, the JMDC database primarily includes working-age individuals, limiting its generalizability to older CKD populations. Finally, the model relied on baseline data and did not incorporate longitudinal trajectories, which may have improved the prediction. However, the model demonstrated consistent risk stratification and clinically coherent predictors using routinely available baseline information, supporting its potential role as an early risk identification tool in real-world settings.

Initiation of emergency dialysis represents a distinct and clinically important outcome that is not fully captured by conventional kidney failure prediction models. In this study, a random forest model based on baseline data achieved moderate discrimination and identified clinically plausible predictors of emergency initiation. Although the model appears to be most useful for early risk stratification, it provides a framework for identifying patients at risk of suboptimal dialysis transition. Future studies should focus on external validation and integration of predictions with structured care pathways to reduce initiation of emergency dialysis in practice.

## Authors’ contributions

KH conceptualized and administered the project, curated and validated the data, conducted formal analysis, devised the methodology, oversaw resource acquisition, visualized the results, and drafted and reviewed the manuscript. TS contributed to the methodology, data interpretation, and critical revision of the manuscript. AW assisted with validation, visualization, and manuscript review. KK assisted with the data curation and data interpretation. YK supervised the study. All authors discussed the results and implications, commented on the manuscript at all stages, and approved the final version.

## Disclosure

The authors declare no conflicts of interest.

## Ethics approval

This study was approved by the Ethics Committee of the University of Tokyo Hospital (Approval No. 2024103NIe) and was conducted in accordance with the Declaration of Helsinki.

## Consent to participate

We used an anonymized dataset in compliance with the protocol and conducted the study using an opt-out approach.

## Consent to publish

Not applicable.

## Data availability statement

The data used in this study cannot be shared publicly because of privacy restrictions. De-identified analytical details may be made available from the corresponding author upon reasonable request, subject to data use restrictions.

