## Supplementary materials for "Development and Validation of Machine Learning Models for Predicting Initiation of Emergency Dialysis in Advanced Chronic Kidney Disease"

**Table of Contents**

**Supplementary Table 1.** Discriminative performance of machine learning models in
the derivation and validation cohorts

**Supplementary Table 1.** Discriminative performance of machine learning models in the derivation and validation cohort

A.	Derivation cohort

|  | LR | RF | SVM | LGBM | XGB |
| --- | --- | --- | --- | --- | --- |
| Accuracy | 0.693  (0.676–0.713) | 0.674  (0.655–0.693) | 0.632  (0.614–0.650) | 0.619  (0.602–0.637) | 0.592  (0.571–0.612) |
| Precision | 0.161  (0.136–0.188) | 0.154  (0.130–0.177) | 0.141  (0.121–0.161) | 0.150  (0.130–0.172) | 0.139  (0.119–0.158) |
| Recall | 0.730  (0.668–0.789) | 0.741  (0.675–0.805) | 0.762  (0.697–0.821) | 0.865  (0.810–0.910) | 0.849  (0.796–0.897) |
| F1score | 0.264  (0.227–0.302) | 0.255  (0.221–0.289) | 0.238  (0.208–0.268) | 0.256  (0.224–0.287) | 0.239  (0.209–0.267) |
| ROC_AUC | 0.749  (0.719–0.780) | 0.752  (0.721–0.785) | 0.736  (0.704–0.768) | 0.774  (0.744–0.800) | 0.745  (0.713–0.776) |
| PR_AUC | 0.180  (0.147–0.233) | 0.170  (0.141–0.216) | 0.171  (0.139–0.218) | 0.178  (0.150–0.220) | 0.172  (0.137–0.221) |
| MCC | 0.234  (0.193–0.273) | 0.225  (0.185–0.265) | 0.206  (0.169–0.241) | 0.247  (0.214–0.276) | 0.222  (0.188–0.251) |

Performance metrics included accuracy, precision, recall, F1 score, C-statistics, PR-AUC, and Matthew’s correlation coefficient. Abbreviations: LR, logistic regression; RF, random forest; SVM, support vector machine; LGBM, Light Gradient Boosting Machine; XGB, extreme gradient boosting; PR-AUC, area under the precision-recall curve; MCC, Matthew correlation coefficient.

B.	Test cohort

|  | LR | RF | SVM | LGBM | XGB |
| --- | --- | --- | --- | --- | --- |
| Accuracy | 0.693  (0.656–0.732) | 0.688  (0.651–0.724) | 0.654  (0.615–0.690) | 0.679  (0.644–0.715) | 0.604  (0.563–0.643) |
| Precision | 0.179  (0.130–0.238) | 0.180  (0.130–0.235) | 0.169  (0.122–0.217) | 0.175  (0.126–0.230) | 0.165  (0.123-0.210) |
| Recall | 0.731  (0.605–0.862) | 0.750  (0.625–0.865) | 0.788  (0.674–0.894) | 0.750  (0.640–0.871) | 0.904  (0.820–0.980) |
| F1score | 0.288  (0.217–0.362) | 0.290  (0.218–0.364) | 0.279  (0.209–0.346) | 0.284  (0.211–0.358) | 0.279  (0.216–0.342) |
| ROC_AUC | 0.792  (0.736–0.851) | 0.799  (0.740–0.850) | 0.781  (0.721–0.842) | 0.794  (0.742–0.844) | 0.781  (0.727–0.830) |
| PR_AUC | 0.261  (0.180–0.377) | 0.242  (0.161–0.354) | 0.255  (0.174–0.379) | 0.237  (0.165–0.370) | 0.206  (0.144–0.305) |
| MCC | 0.246  (0.168–0.329) | 0.252  (0.172–0.330) | 0.245  (0.173–0.316) | 0.244  (0.170–0.314) | 0.268  (0.205–0.326) |

Performance metrics included accuracy, precision, recall, F1 score, C-statistics, PR-AUC, and Matthew’s correlation coefficient. Abbreviations: LR, logistic regression; RF, random forest; SVM, support vector machine; LGBM, Light Gradient Boosting Machine; XGB, extreme gradient boosting; PR-AUC, area under the precision-recall curve; MCC, Matthew correlation coefficient.
